# Loss-of-Function *FLNC* Variants are Associated with Arrhythmogenic Cardiomyopathy Phenotypes when Identified through Exome Sequencing of a General Clinical Population

**DOI:** 10.1101/2021.10.28.21265491

**Authors:** Eric D. Carruth, Maria Qureshi, Amro Alsaid, Melissa A. Kelly, Hugh Calkins, Brittney Murray, Crystal Tichnell, Amy C. Sturm, Regeneron Genetics Center, Aris Baras, H. Lester Kirchner, Brandon K. Fornwalt, Cynthia A. James, Christopher M. Haggerty

## Abstract

**Background:** The *FLNC* gene has recently garnered attention as a likely cause of arrhythmogenic cardiomyopathy (ACM), which is considered an actionable genetic condition. However, the association with disease in an unselected clinical population is unknown. We hypothesized that individuals with loss-of-function variants in *FLNC* (*FLNC*_LOF_) would have increased odds for ACM-associated phenotypes versus variant-negative controls in the Geisinger MyCode cohort.

**Methods:** We identified rare, putative *FLNC*_LOF_ among 171,948 individuals with exome sequencing linked to health records. Associations with ACM phenotypes from available diagnoses and cardiac evaluations were investigated.

**Results:** Sixty individuals (0.03%; median age 58 years [47–70 IQR], 43% male) harbored 27 unique *FLNC*_LOF_. These individuals had significantly increased odds ratios (OR) for dilated cardiomyopathy (OR:4.9, [95% confidence interval: 2.6–7.6]; p<0.001), supraventricular tachycardia (OR:3.2, [1.1–5.6]; p=0.01), defibrillator implantation (OR:4.6, [1.9–8.4]; p<0.001), and left-dominant ACM (OR:4.2, [1.4–7.9]; p=0.003). Echocardiography revealed reduced left ventricular ejection fraction (52±13% vs. 57±9%; p=0.001) associated with *FLNC*_LOF_. Overall, at least 9% of *FLNC*_LOF_ carriers demonstrated evidence of penetrant disease.

**Conclusions:** *FLNC*_LOF_ variants are associated with increased odds of ventricular arrhythmia and dysfunction in an unselected clinical population. These findings support genomic screening of *FLNC* for actionable secondary findings.

## INTRODUCTION

The *FLNC* gene, which encodes the filamin-C sarcomeric protein, has recently garnered attention as a potential cause of arrhythmogenic cardiomyopathy (ACM).^1^ Early reports have shown convincing associations of *FLNC* variants with an overlapping phenotype of dilated cardiomyopathy (DCM) and ACM, which is typically left-dominant (LDAC).^2–5^ Commonly cited characteristics from clinical evaluation of individuals with *FLNC* variants have included frequent ventricular arrhythmias or sudden cardiac death, reduced left ventricular (LV) systolic function, and regional structural abnormalities such as non-ischemic late gadolinium enhancement (LGE) patterns on cardiac MRI (CMR).^6–10^

To date, these studies have primarily investigated gene-phenotype associations in individuals or families with known cardiomyopathy. However, little is known of the disease burden and phenotype characteristics associated with *FLNC* variants when identified through broad exome sequencing within an unselected clinical population. Such data are critical for understanding the disease penetrance of these variants when discovered as a secondary or incidental finding, and to thus guide clinical management of patient-carriers. This consideration is particularly timely given the recent inclusion of *FLNC* in the American College of Medical Genetics and Genomics (ACMG) recommendation for clinical return of secondary findings.^11^

The advent of large genomic screening initiatives provides opportunities to identify *FLNC* variants—and characterize the associated phenotype—in a relatively unselected population. With one of the largest sequenced biobanks linked to a long-standing electronic health records (EHR) system, Geisinger’s MyCode Community Health Initiative enables such an assessment. Using this resource, the phenotypic burden in individuals with rare, putative loss-of-function *FLNC* variants (*FLNC*_LOF_) was retrospectively investigated and compared to the remaining sequenced MyCode cohort using EHR-based phenotypes. Additionally, blinded manual chart review of available cardiac evaluations in these individuals was performed and compared with matched controls. We hypothesized that MyCode participants with *FLNC*_LOF_ would have increased odds for ACM-associated phenotypes compared to controls.

## PATIENTS AND METHODS

### Study Information

Geisinger’s MyCode Community Health Initiative [2007–Present] is an IRB-approved research and precision health project with over 290,000 participants and counting. Opt-in informed consent is obtained for collection of biospecimens and linkage to EHR for broad research use.^12^ Through the DiscovEHR collaboration with Regeneron Genetics Center (RGC; Tarrytown, NY), these biospecimens (participants’ blood or saliva) are used to extract genomic DNA and generate exome sequence data. The first 175,509 sequenced MyCode participants were included in this study.

### Exome Sequencing, Variant Calling, and Genotype Assignment

The sequencing methodology followed standard practices at the RGC, which have been described in detail previously.^13–15^ Briefly, probes from Nimblegen (VCRome, referred to as VCR henceforth) or a version of the xGEN probe from Integrated DNA Technologies (IDT) were used for target sequence capture. Sequencing was performed by paired end 75bp reads on either an Illumina HiSeq2500 or NovaSeq. Coverage depth was sufficient to provide more than 20% coverage over 85% of the targeted bases in 96% of the VCR samples and 90% coverage for 99% of IDT samples.

Following sequencing, samples showing disagreement between genetically determined and reported sex, low quality sequence data, genetically-identified sample duplicates, and samples lacking linked EHR data were excluded. After these exclusions, 171,948 participants were retained for analysis.

Alignments and variant calling were based on GRCh38 human genome reference sequence. Variant calls were produced using the WeCall variant caller (https://github.com/Genomicsplc/wecall; version 1.1.2). A project-level VCF (pVCF) was compiled using the GLnexus joint genotyping tool (version 1.1.3-4).^16^

### *FLNC*_LOF_ Variant Annotation and Selection

All rare (MAF < 0.001) variants in *FLNC* were annotated using Ensembl VEP to identify ‘High’ impact, putative loss-of-function variants (frameshift, stop-gain, and canonical splice site) exclusive of those in the terminal exon.^17^ Sample-level variant calls with site read depth < 7, and alternate allele balance < 20% (or fewer than 5 alternate reads) were removed.

### Phenotype Characterization

#### EHR-based Comparison

Available demographics (age at last encounter, sex, body mass index) and diagnosis codes (International Classification of Disease, 9^th^ or 10^th^ edition (ICD9, ICD10), as applicable) from 1996–Present were retrieved for all *FLNC*_LOF_ individuals and all variant-negative controls who lacked any rare LOF or non-synonymous substitution variants (not classified as ‘benign’ in ClinVar) in a broad panel of cardiomyopathy- or arrhythmia-associated genes (Supplementary Table 2). Additionally, structured tabular data from echocardiogram and Holter monitor studies were gathered for all patients, as available (taking only the most recent if multiple existed).

Phenotypes for dilated cardiomyopathy (DCM), ventricular tachycardia (VT), ventricular fibrillation (VF), supraventricular tachycardia (SVT), premature beats (including premature ventricular contractions), cardiac arrest, and implantable cardioverter-defibrillator (ICD) placement were defined based on the presence of ICD9/ICD10 codes on patient problem lists or ≥ 2 encounter diagnoses (except VF and cardiac arrest, for which a single encounter diagnosis was accepted). Phenotypes for heart failure (HF) and atrial fibrillation (AF) were defined by internally developed and previously validated algorithms, as described in the Online Supplement.^18,19^

Finally, based on prior work,^20^ a composite phenotype for LDAC was defined as comprising histories of both ventricular arrhythmia and LV dysfunction, as follows:

1. Arrhythmia (either of the following):
  a. Premature ventricular contractions (PVCs) in >0.5% of QRS complexes on Holter monitor
  b. Any diagnoses of VT, VF, SVT, premature beats, or cardiac arrest
2. LV dysfunction (either of the following):
  a. LV ejection fraction (LVEF) <50% on most recent echocardiogram
  b. Past diagnosis of heart failure (without ischemic cardiomyopathy) or other non-ischemic cardiomyopathy

The prevalence of this LDAC classification and its components were compared between *FLNC*_LOF_ and the remaining MyCode cohort based on the above phenotypes.

#### Physician Chart Review

All individuals with *FLNC*_LOF_ variants were 2-to-1 matched by age and sex to controls drawn from the broader control group, as described above. Dual manual chart review was performed by two experts (MQ and AA) on this combined set (blinded to group assignment) to further characterize histories of ventricular dysfunction and arrhythmic events such as ventricular tachycardia and cardiac arrest. Additionally, the most recent interpretable ECG tracing for each patient was reviewed by a single expert (HC) for the diagnostic 2010 arrhythmogenic right ventricular cardiomyopathy (ARVC) task force criteria (TFC).^21^

The presence or absence of ACM was defined during this chart review based on the 2019 Heart Rhythm Society’s consensus statement.^22^ In brief, ACM is defined by ventricular dysfunction unexplained by ischemia, hypertension, or valvular disease, where arrhythmia is part of the presentation of the dysfunction. Only systolic dysfunction was included, and arrhythmias were included only if observed within 1 year of the presentation of dysfunction.

### Statistics

Data are presented as mean ± standard deviation, median [inter-quartile range (IQR)], or count (%), as appropriate. To compare the prevalence of diagnoses of interest between *FLNC*_LOF_-positive individuals and the rest of the sequenced cohort, Firth’s bias-reduced logistic regression was used (implemented using the ‘logistf’ package in R).^23^ To account for population relatedness, a bootstrap procedure (1000 iterations) was used to estimate the variance in the regression coefficient. Similarly, bootstrapped linear regressions with 1000 iterations were used to evaluate associations of quantitative echocardiography measures with *FLNC*_LOF_ genotype. All models were adjusted for age, sex, and the first four principal components of ancestry. For all group comparisons based on chart reviews, categorical data were compared using Fisher’s exact test. False discovery rate correction was applied as per Benjamini-Hochberg.^24^ Group differences of categorical data were considered statistically significant if the adjusted p<0.05 and the 95% confidence interval of the odds ratio did not cross one.

## RESULTS

A total of 171,948 MyCode participants had available sequencing data linked to their EHR. Of these, 60 individuals (0.03%) were identified harboring 27 unique *FLNC*_LOF_ variants, comprising frameshift (n=14), splice site (10), and stop gained (3). Variant details are presented in Table 1. Basic demographics of *FLNC*_LOF_ individuals were comparable to the remaining MyCode cohort lacking a LOF *FLNC* variant (Table 2).

**Table 1.** *FLNC*_LOF_ variant details in the MyCode cohort.

| CHR:POS | HGVSc<br>(NM_001458.5) | HGVSp<br>(NP_001449.3) | Consequence | ClinVar<br>Review<br>Status | N Pts |
| --- | --- | --- | --- | --- | --- |
| 7:128835324 | c.353-2A>G | -- | Splice | . | 1 |
| 7:128837255 | c.697C>T | p.Gln233* | Nonsense | P, 1-star | 1 |
| 7:128841281 | c.1926_1947del | p.Ile643Glufs*21 | Frameshift | . | 2 |
| 7:128841364 | c.2007+1G>A | -- | Splice | . | 1 |
| 7:128841529 | c.2084del | p.Arg695Leufs*8 | Frameshift | P, 1-star | 1 |
| 7:128841565 | c.2119C>T | p.Gln707* | Nonsense | P, 1-star | 7 |
| 7:128842375 | c.2265+1G>A | -- | Splice | LP, 1-star | 4 |
| 7:128842375 | c.2265+1G>T | -- | Splice | LP, 1-star | 4 |
| 7:128843418 | c.2652_2653insG | p.Lys886Glufs*34 | Frameshift | . | 2 |
| 7:128843861 | c.2878del | p.Val960Trpfs*29 | Frameshift | . | 1 |
| 7:128844656 | c.3193-2A>G | -- | Splice | LP, 1-star | 2 |
| 7:128845989 | c.3791-1G>C | -- | Splice | P/LP, 2-star | 2 |
| 7:128846094 | c.3901_3904del | p.Asp1301Profs*3 | Frameshift | . | 1 |
| 7:128846744 | c.4128-1G>C | -- | Splice | . | 8 |
| 7:128848974 | c.4926_4927insACGTCAC<br>A | p.Val1643Thrfs*26 | Frameshift | P, 1-star | 1 |
| 7:128849329 | c.4952-2A>T | -- | Splice | LP, 1-star | 1 |
| 7:128851361 | c.5668+1G>C | -- | Splice | . | 4 |
| 7:128851539 | c.5754del | p.Leu1919Cysfs*34 | Frameshift | . | 1 |
| 7:128852730 | c.5983del | p.Arg1995Alafs*21 | Frameshift | P, 1-star | 1 |
| 7:128853517 | c.6258del | p.Asn2087Thrfs*40 | Frameshift | . | 1 |
| 7:128854163 | c.6676del | p.Leu2226Trpfs*25 | Frameshift | . | 1 |
| 7:128855206 | c.7143T>G | p.Tyr2381* | Nonsense | . | 2 |
| 7:128856630 | c.7365del | p.Tyr2455* | Nonsense | LP, 1-star | 4 |
| 7:128856636 | c.7371del | p.Glu2458Serfs*71 | Frameshift | P, 1-star | 4 |
| 7:128856856 | c.7496_7497insTGCT | p.Gln2499Hisfs*46 | Frameshift | P, 1-star | 1 |
| 7:128857312 | c.7757del | p.Ser2586Thrfs*29 | Frameshift | . | 1 |
| 7:128858006 | c.7781-2A>C | -- | Splice | . | 1 |

Coordinates are with respect to GRCh38. ClinVar review as of April 2021. P:
pathogenic; LP: likely pathogenic.

**Table 2.**
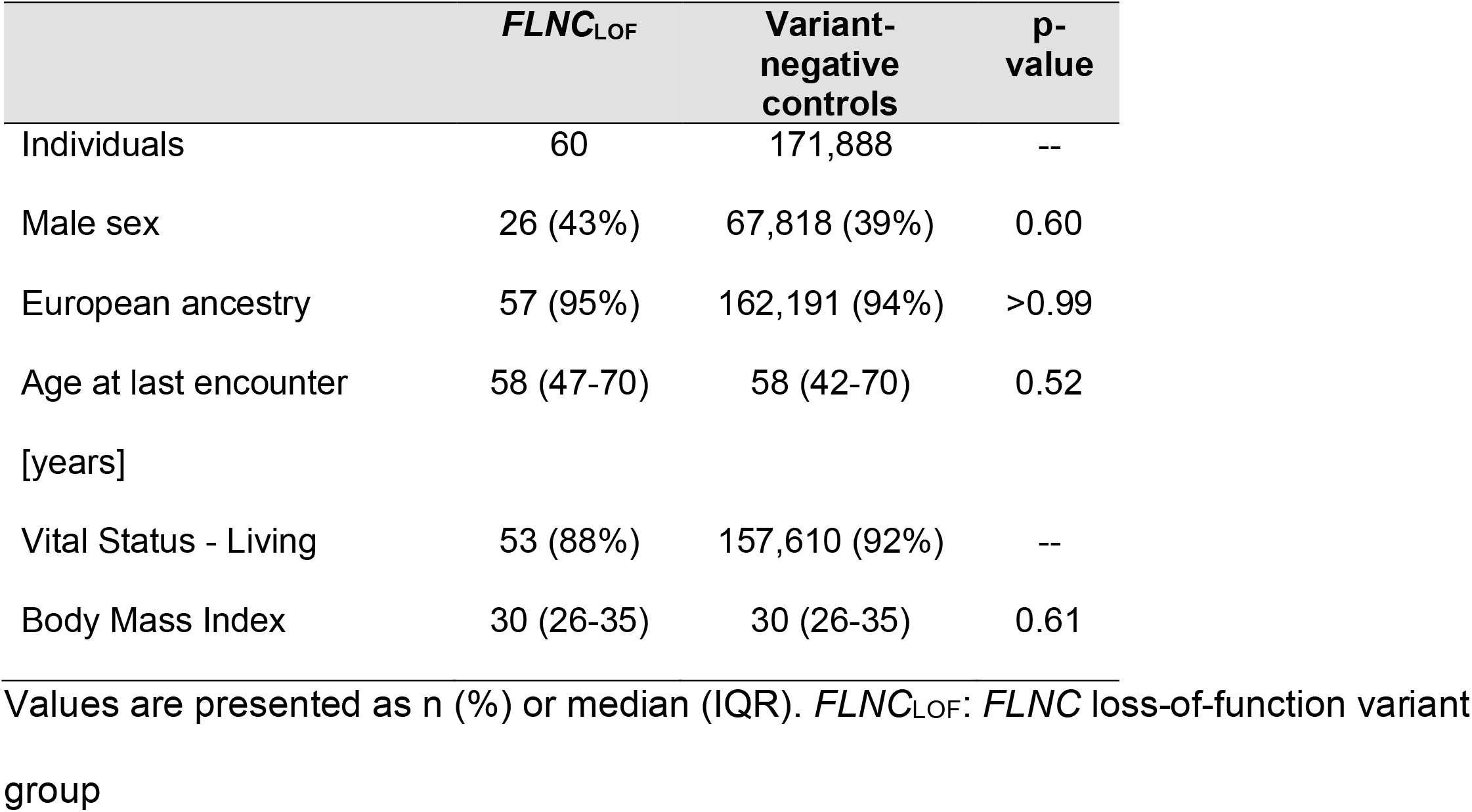
Demographics of *FLNC*_LOF_ group vs. variant-negative controls in MyCode.

|  | *FLNC*<sub>LOF</sub> | Variant-negative controls | p-value |
| --- | --- | --- | --- |
| Individuals | 60 | 171,888 | -- |
| Male sex | 26 (43%) | 67,818 (39%) | 0.60 |
| European ancestry | 57 (95%) | 162,191 (94%) | >0.99 |
| Age at last encounter<br>[years] | 58 (47-70) | 58 (42-70) | 0.52 |
| Vital Status - Living | 53 (88%) | 157,610 (92%) | -- |
| Body Mass Index | 30 (26-35) | 30 (26-35) | 0.61 |

Values are presented as n (%) or median (IQR). *FLNC*<sub>LOF</sub>: *FLNC* loss-of-function variant group

### *FLNC*_LOF_ are associated with increased odds of disease and ventricular remodeling/dysfunction diagnoses in the EHR

Comparing the EHR-derived phenotypes of individuals with *FLNC*_LOF_ to the rest of the MyCode cohort, *FLNC*_LOF_ was associated with significantly greater odds ratios (OR) of heart disease diagnoses, including DCM (OR:4.9 [95% CI 2.6–7.6]), SVT (OR:3.2 [1.1-5.6]), and ICD (OR:4.6 [1.9–8.4]), as shown in Table 3. Ventricular dysfunction (with or without arrhythmia) was observed in 15/60 (25%) *FLNC*_LOF_ individuals vs. 19,461/171,888 (11%) variant-negative controls (OR:3.0 [1.6–5.0]; p<0.001). Arrhythmia was present in 9/60 (15%) *FLNC*_LOF_ vs. 13,808/171,888 (8%) controls (OR:2.1 [0.9–3.6]; p=0.06). Together, 6/60 (10%) *FLNC*_LOF_ carriers had history of both ventricular dysfunction and arrhythmia—suggestive of LDAC—as compared to 4,903/171,888 (3%) of the remaining variant-negative cohort (OR:4.2 [1.4–7.9]; p=0.003; Table 3; Figure 1A).

**Table 3.** Associations of cardiomyopathy phenotypes in *FLNC*_LOF_ group vs. variant-negative controls in MyCode.

| Phenotype | *FLNC*<sub>LOF</sub><br>(n=60) | Variant-negative<br>controls<br>(n=171,888) | OR | 95% CI | *p-value |
| --- | --- | --- | --- | --- | --- |
| DCM | 9 (15%) | 6,341 (4%) | 4.9 | 2.6 – 7.6 | 1.7e-7 |
| HF | 9 (15%) | 14,333 (8%) | 2.2 | 0.96 – 3.90 | 0.04 |
| CM | 9 (15%) | 6,890 (4%) | 4.5 | 2.4 – 7.0 | 4.7e-7 |
| AF | 7 (12%) | 16,408 (10%) | 1.3 | 0.6 – 2.3 | 0.51 |
| SVT | 6 (10%) | 6,171 (4%) | 3.2 | 1.1 – 5.6 | 0.01 |
| PVCs | 1 (2%) | 3,828 (2%) | 1.1 | 0.5 – 2.1 | 0.91 |
| ICD | 4 (7%) | 2,907 (2%) | 4.6 | 1.9 – 8.4 | 8.7e-4 |
| LDAC | 6 (10%) | 4,903 (3%) | 4.2 | 1.4 – 7.9 | 0.003 |

*FLNC*<sub>LOF</sub>: *FLNC* loss-of-function variant group; OR: odds ratio; CI: confidence interval; DCM: dilated cardiomyopathy; HF: heart failure; CM: Cardiomyopathy; AF: atrial fibrillation; VT: ventricular tachycardia; VF: ventricular fibrillation; SVT: supraventricular tachycardia; PVCs: premature ventricular contractions; ICD: implantable cardioverter defibrillator; LDAC: left-dominant arrhythmogenic cardiomyopathy. \*p-values based on Firth's logistic regression model adjusted for age, sex, and ancestry and adjusted for multiple comparisons (p<0.05 and OR 95% CI not crossing 1 considered significant).

**Figure 1.**
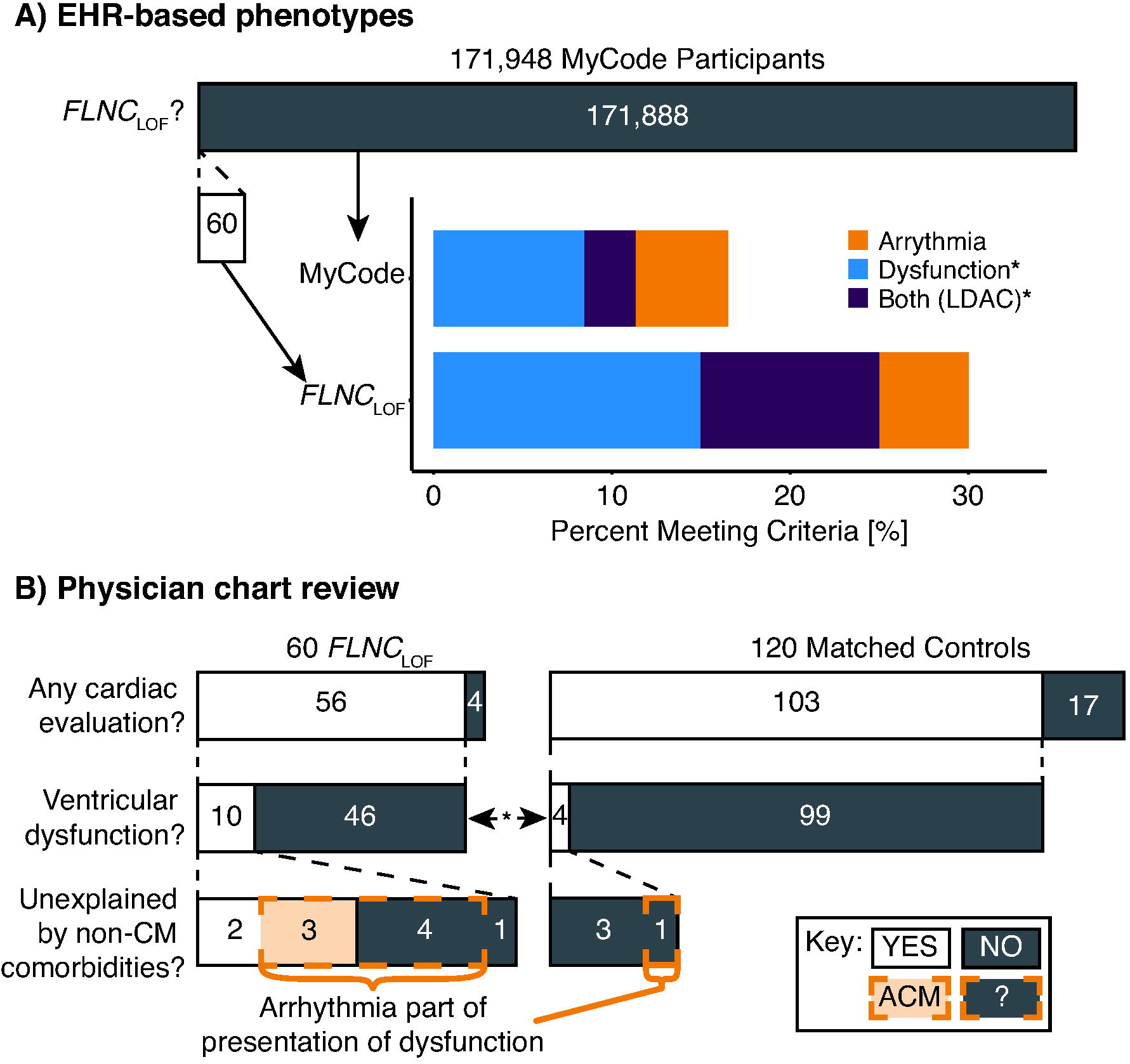
Summary of phenotypic findings in *FLNC*_LOF_ individuals. (A) Compared to the rest of the MyCode population, individuals with *FLNC*_LOF_ were significantly enriched for EHR-based phenotypes of ventricular dysfunction (p<0.001) and both dysfunction and arrhythmia (LDAC; p=0.003), but not for arrhythmia criteria (p=0.06). (B) ACM 2019 criteria from matched chart review revealed that individuals with *FLNC*_LOF_ were enriched for ventricular dysfunction (p=0.04). Similar apparent enrichment was observed for unexplained dysfunction and having arrhythmia as part of the presentation of the dysfunction, however these group differences were not significant. EHR: electronic health records; LDAC: left-dominant arrhythmogenic cardiomyopathy; *FLNC*_LOF_: *FLNC* loss-of-function variant group; CM: cardiomyopathy; ACM: arrhythmogenic cardiomyopathy-associated phenotype; ?: represents those with dysfunction confounded by other co-morbidities and arrhythmia, not classified as ACM. *p<0.05 for groupwise comparisons.

In addition to the diagnostic data, echocardiograms were available for 32/60 (53%) *FLNC*_LOF_ individuals and 75,837/171,888 (44%) variant-negative controls (Table 4). Those in the *FLNC*_LOF_ group had significantly reduced LVEF (52 ± 13% *FLNC*_LOF_ vs. 57 ± 9% controls; p=0.001; Figure 2) and increased left ventricular end-diastolic inner diameter (LVIDd; 4.9 ± 0.8 cm *FLNC*_LOF_ vs. 4.6 ± 0.7 cm; p=0.02) compared to controls. However, the indexed LVIDd was not significantly different (2.5 ± 0.4 cm/m^2^ *FLNC*_LOF_ vs. 2.4 ± 0.4 cm/m^2^; p=0.10) Interventricular septum thickness in diastole (IVSd) was also slightly increased (1.12 ± 0.21 cm *FLNC*_LOF_ vs. 1.08 ± 0.24 cm controls; p=0.02).

**Table 4.** Associations of echocardiogram measures with *FLNC*_LOF_.

| **Echocardiogram**<br>**measure** | ***FLNC*<sub>LOF</sub>** | **Variant-negative control** | **p-value\*** |
| --- | --- | --- | --- |
| Individuals with available<br>studies (% of group) | 32 (53%) | 75,837 (44%) |  |
| LVEF [%] | 52 ± 13 | 57 ± 9 | 0.001 |
| LVIDd [cm] | 4.9 ± 0.8 | 4.6 ± 0.7 | 0.02 |
| IVSd [cm] | 1.12 ± 0.21 | 1.08 ± 0.24 | 0.02 |

*FLNC*<sub>LOF</sub>: *FLNC* loss-of-function variant group; LVIDd: left ventricular internal diameter and end-diastole; IVSd: interventricular septum dimension at end-diastole; LVEF: left ventricular ejection fraction. \*p-values based on linear model adjusted for age, sex, and ancestry and adjusted for multiple comparisons (p<0.05 considered significant).

**Figure 2.**
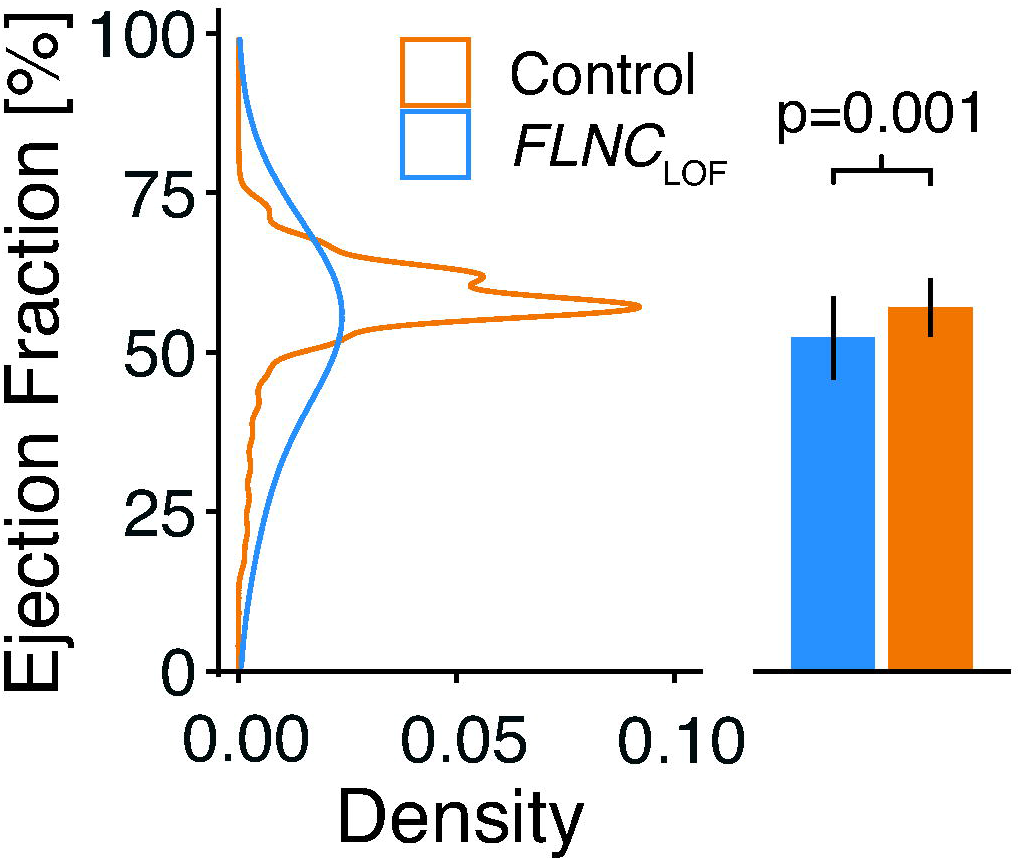
Most recent left ventricular ejection fraction in individuals with *FLNC*_LOF_ variants and variant-negative controls. Ejection fraction was significantly reduced in the *FLNC*_LOF_ group compared to controls, as observed using a bootstrapped linear regression model (p=0.001). *FLNC*_LOF_: *FLNC* loss-of-function variant group.

### Chart review confirms association with arrhythmias and cardiomyopathies and identification of ACM

Results from matched chart reviews are summarized in Table 5. Overall, the burden of cardiomyopathy, ventricular tachycardia (VT), and sudden cardiac arrest (SCA) was observed to be higher (OR >3) in the *FLNC*_LOF_ group than matched controls—consistent with the EHR-based analysis—but these observations were not independently statistically significant in this smaller sample.

**Table 5.**
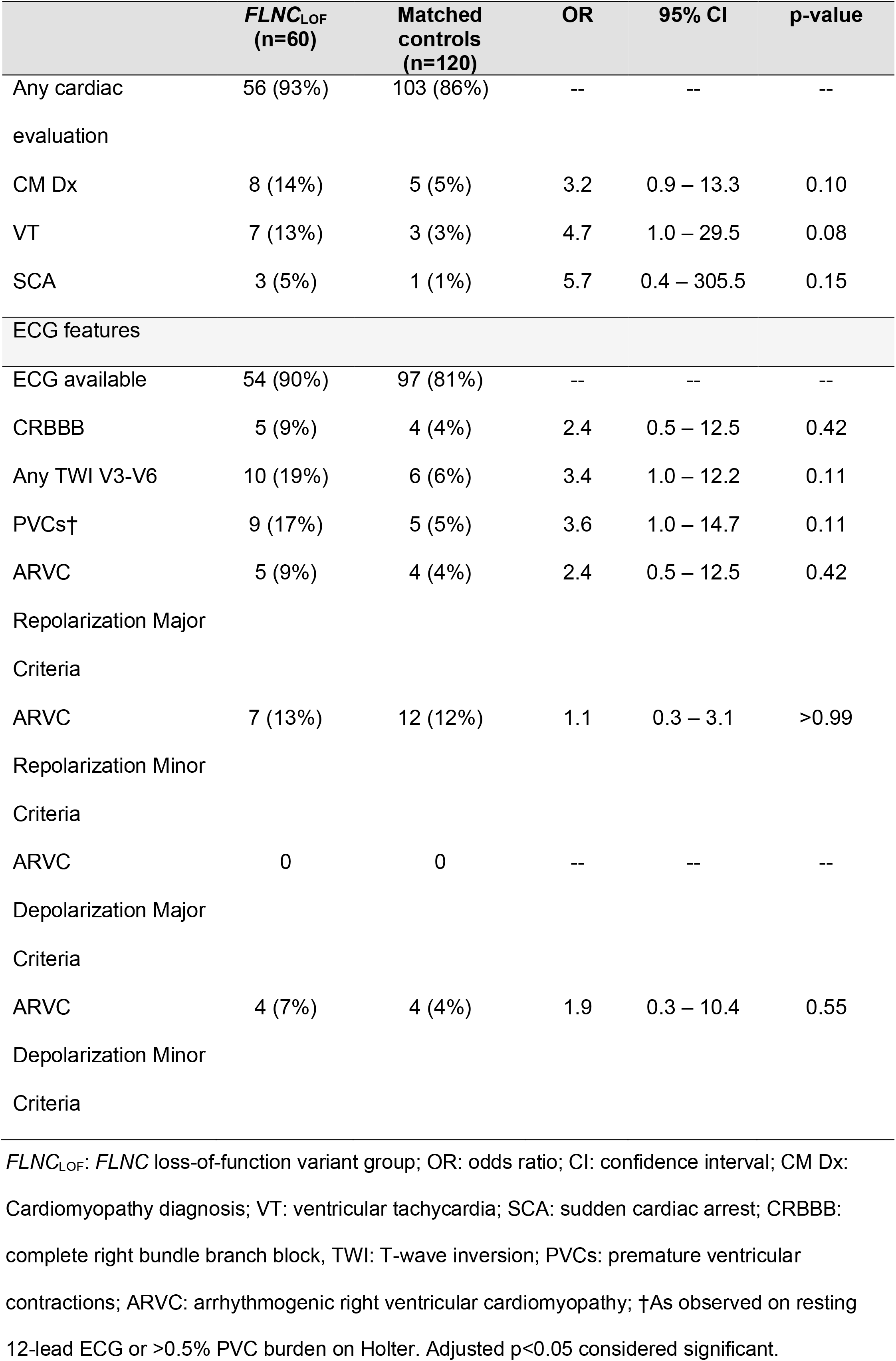
Results of manual chart review in *FLNC*_LOF_ group and matched controls.

The most recent non-paced, interpretable ECG was evaluated for each patient (54/60 available for *FLNC*_LOF_ group and 97/120 for matched controls) to identify the presence of any depolarization, repolarization, or arrhythmia abnormalities, such as those commonly used to diagnose ARVC. ARVC-specific TFC were uncommon (4– 13%) and comparable in frequency between groups (Table 5). Other characteristics, such as T-wave inversions in any anterior or lateral precordial leads (V3-V6) and PVCs on ECG or Holter (>0.5% burden) were observed more frequently (OR >3) in the *FLNC*_LOF_ group, but those differences were not statistically significant (Table 5; p=0.11 for both).

Finally, a detailed evaluation of the presence, form, and etiology of any cardiomyopathy present in the subset of these patients with any cardiac evaluation (56/60 *FLNC*_LOF_ and 103/120 matched controls) was performed. Ventricular dysfunction was present in 10/56 (18%) *FLNC*_LOF_ individuals vs. 4/103 (4%) matched controls (OR:5.3 [1.4–24.5]; p=0.04; Figure 1B), and arrhythmia was part of the presentation of the dysfunction for seven of these patients in the *FLNC*_LOF_ group (7/10; 70%) vs. 1/4 (25%) controls (OR:6.0 [0.3–417.2]; p=0.24). Moreover, for 50% of the 10 *FLNC*_LOF_ carriers with dysfunction (5/56 (9%) overall), the ventricular dysfunction was unexplained by ischemia, hypertension, or valvular disease compared with 0/4 controls with dysfunction (p=0.24). Thus, by strict application of the 2019 consensus statement, 3/56 *FLNC*_LOF_ individuals (5%) satisfied the criteria for classification as ACM vs. 0/103 controls (p=0.10).

The three individuals identified as having an ACM-associated phenotype had the following characteristics. All three were male. One was diagnosed with DCM in his 40s, with concurrent ventricular tachycardia and resultant syncope. An ICD was placed in this patient for secondary prevention (with a subsequent appropriate shock), and the pathogenic *FLNC* variant in this individual was known via clinical genetic testing. Another individual had New York Heart Association class II HF in his 70s with AF and frequent PVCs noted on ECG, with an ICD placed for primary prevention. The third (in his 70s) had a globally hypokinetic LV, with atypical late gadolinium enhancement in the midwall on CMR, with non-sustained VT and polymorphic PVCs.

Of the remaining two *FLNC*_LOF_ individuals with unexplained ventricular dysfunction (and no concomitant arrhythmia), one was a female diagnosed with DCM in her 40s, with later development of non-sustained VT, ICD and left ventricular assist device placement. The likely pathogenic *FLNC* variant was known via clinical genetic testing in this individual. The other was also female, with normal LVEF but diffuse right ventricular hypokinesis in her 50s, confounded by lung cancer with chronic malignant pericardial effusion. This patient also later developed supraventricular tachycardia and AF, with frequent PVCs noted on ECG.

## DISCUSSION

Genetic variants associated with ‘high-risk’ arrhythmogenic conditions—such as pathogenic variants in *LMNA, PKP2*, and *SCN5A*—are increasingly being considered medically actionable and recommended for clinical genomic screening.^11^ Loss-of-function variants in *FLNC* have been recognized as a potential cause of DCM/ACM and heart failure from disease-based cohorts,^2,7,9,25^ and indeed, this recognition has led to the recent addition of *FLNC* to such professional society recommendations for secondary genomic findings.^11^ However, the disease burden associated with *FLNC* variants in broader clinical populations (i.e., not ascertained through clinical symptoms or family history) have not been described to inform clinical management in relation to such secondary findings. This work begins to address this gap by evaluating the prevalence and associated phenotype of individuals with *FLNC*_LOF_ variants in the MyCode cohort—a large, healthcare-seeking population. In this cohort, individuals carrying *FLNC*_LOF_ variants have increased odds for heart disease-associated phenotypes (cardiomyopathy and arrhythmia) as well as quantitative changes in cardiac structure and function compared with controls. Moreover, detailed cardiologist chart review confirmed a significantly increased burden of ventricular dysfunction associated with *FLNC*_LOF_, half of which was not explained by ischemia, hypertension, or valve disease; and with a large proportion (70%) presenting with concurrent arrhythmia. In contrast, few individuals had ventricular dysfunction in age- and sex-matched controls, all of which was explained by ischemia, hypertension, or valve disease, and only one of which (25%) had concurrent arrhythmia. Furthermore, the remaining two with unexplained dysfunction eventually developed arrhythmia. These data provide strong evidence that incidental/secondary identification of *FLNC*_LOF_ is both medically relevant and actionable via medication, arrhythmia monitoring, or if warranted, ICD placement.

### Phenotype Characteristics of *FLNC*_LOF_ through Genome-First Approach

There has been considerable attention over the last 5 years to defining the disease characteristics associated with *FLNC* variants identified in large disease-based cohorts. These studies have broadly found that cardiomyopathy in individuals with *FLNC*_LOF_ is characterized by LV dysfunction/dilation, a high burden of malignant arrhythmia/sudden cardiac arrest, frequent findings of myocardial fibrosis, and reported high rates of penetrance.^2,3,6,10,26,27^ Despite the stark differences in clinical context, the characteristics of this genome-first cohort generally recapitulate these findings. For example, *FLNC*_LOF_ variants were associated with LV dilatation (increased LVIDd) and decreased LVEF, with 25% of carriers being classified as having ventricular dysfunction from EHR-based phenotypes (18% via chart review). Similarly, while affecting relatively small proportions of *FLNC*_LOF_ carriers, this analysis revealed evidence of potentially severe arrhythmias based on significantly increased use of implanted defibrillators, significantly increased odds of SVT, and 13% and 5% of patients with history of VT and sudden cardiac arrest, respectively.

Characterizing the risk of severe arrhythmia for individuals with *FLNC*_LOF_ is particularly important given that *FLNC* is included in the Heart Rhythm Society’s list of genes that warrant consideration for ICD placement for primary prevention of ACM-associated sudden cardiac death.^22^ Furthermore, emerging data suggest that, as for *LMNA* and *DSP*,^28^ significant arrhythmia risk may be present with *FLNC*_LOF_ even in the setting of only mild reductions in LVEF,^10,27^ potentially requiring alternate criteria for use. The application of these criteria to the specific case of secondary findings warrants particular attention and additional study.

In that regard, consideration of the apparent penetrance observed in this cohort is relevant. While prior studies have reported disease penetrance as high as 70% in *FLNC*_LOF_ families with known cardiomyopathy,^2^ the proportion of individuals with *FLNC*_LOF_ from MyCode exhibiting penetrant disease is considerably lower, which is consistent with similar genome-first observations in other cardiac-associated genes.^15,29,30^ Specifically, 9% of patients had ventricular dysfunction not explained by ischemia, hypertension, or valve disease—arguably representing a conservative estimate of penetrant disease. For instance, it is worth noting that, in an additional 7% of cases, arrhythmia was part of the disease presentation of dysfunction potentially attributable to other co-morbidities. In such cases, the potential modulating effect of the variant on the disease course in those individuals is difficult to quantify. Furthermore, this estimate of penetrance is based on disease history to date, whereas some individuals may yet develop disease during their lifetime. Indeed, of those in the *FLNC*_LOF_ group, affected individuals were apparently older (current median age 71 years [IQR:55-73]) than those unaffected (median 58 [45-71]; p=0.09). Despite this challenge, the observed penetrance estimate is comparable, if not higher, than prior analyses of other cardiomyopathy-associated genes in the MyCode cohort, such as the genes for ARVC (6%)^15^ and *TTN* (5–12%).^31,32^

### *FLNC* as an ACM-Associated Gene

Whereas some studies have reported *FLNC* associations with ARVC;^3,4,6,7,9^ that is, the specific right ventricle-affecting subtype of ACM, the 2010 ARVC TFC exhibited apparently low sensitivity in our population-based cohort, though incomplete testing may have contributed to this low yield. In particular, repolarization and depolarization ECG criteria from the 2010 ARVC TFC were indistinguishable between *FLNC*_LOF_ individuals and controls, a finding consistent with other reports, even in patients meeting other (non-ECG) TFC.^6^ Prospective studies with comprehensive evaluations for TFC will help clarify the prevalence of these and other ARVC TFC in *FLNC*_LOF_.

Instead, this work provides strong evidence that *FLNC*_LOF_ variants primarily lead to left-sided disease. For example, 10% of *FLNC*_LOF_ individuals demonstrated evidence (both ventricular dysfunction and arrhythmia) of LDAC in their EHR. This classification is challenged by the fact that LDAC, as yet, has no formal widely recognized diagnostic criteria.^33^ Some studies have proposed such criteria, which include LV systolic global or regional dysfunction, LGE, evidence of depolarization or repolarization abnormalities on ECG, and evidence of ventricular arrhythmias.^26,34,35^ However, in the current absence of consensus criteria, our definition represents a pragmatic approach that is both conducive to our EHR-based phenotyping and consistent with the broad definition of ACM by current HRS guidelines. These findings also highlight the potential utility of genotype-first approaches to the diagnosis, as well as the management, of ACM.^22^

Of note, several proposals for LDAC criteria have emphasized findings of LGE by CMR in the diagnostic scoring. For example, ring-like LGE patterns in the LV have been preferentially associated with DCM in desmoplakin (*DSP*) and *FLNC* variant carriers.^26,36^ Unfortunately, the availability of contrast-enhanced CMR data was limited in these patients, precluding evaluation of LGE patterns or detailed regional ventricle structure/function, though this finding was observed in at least one *FLNC*_LOF_ individual with ventricular dysfunction. Based on the presumed notion that myocardial fibrosis is the substrate for ventricular arrhythmias in ACM, such an assessment may uncover additional structural evidence of LDAC in *FLNC*_LOF_. Consistent LGE patterns have been reported in multiple disease-based *FLNC* cohorts already.^6,10,26^

### Limitations

This study was from a single healthcare system, representing primarily European ancestry. There is an inherent possibility of survival bias in genomic screening ascertainment, which may have favored unaffected individuals. Variants detected via exome sequencing were not confirmed by orthogonal methods. Only rare, putative loss-of-function variants were investigated herein, so the potential disease association with rare missense variants has not been established and must be investigated. Despite supplementation with cardiologist chart review, the retrospective nature of this study, and the inherent potential for errors or missing data in the EHR, resulted in incomplete testing for ACM phenotypes, particularly Holter monitor data, so there may be additional evidence of disease in either group that was not observed herein. CMR findings were not investigated in this work due to the limited number of studies available, so the prevalence of LGE findings or other structural or functional abnormalities in these individuals is unknown. Finally, the effects of environmental factors, such as exercise history, may contribute to variable disease penetrance, and should be investigated in future studies.

### Conclusion

In this retrospective study, individuals with *FLNC* loss of function variants identified via genome screening had increased odds of DCM and ACM phenotypes, including reduced systolic LV function and common ventricular arrhythmias, compared to variant-negative controls from the MyCode cohort. Physician chart review confirmed these EHR-based associations with cardiomyopathy and arrhythmia, with an estimated disease penetrance of at least 9%. The growing evidence of these associated disease phenotypes in individuals with *FLNC* variants supports the inclusion of this gene for surveillance as part of clinical genomic screening for earlier disease detection and prevention of serious adverse outcomes including sudden cardiac death.

## Data Availability

The data that support the findings of this study will not be made available.

## ACKNOWLEDGMENTS

The authors gratefully acknowledge the participation of MyCode participants and the sequencing efforts of Regeneron Genetics Center.

## AUTHOR INFORMATION

Conceptualization: E.D.C., C.A.J., C.M.H.; Formal analysis: E.D.C., H.L.K., C.M.H.; Funding acquisition: C.A.J., C.M.H.; Investigation: E.D.C., M.Q., A.A., H.C., B.M., C.T., C.M.H.; Methodology: E.D.C., M.Q., A.A., H.L.K., C.M.H.; Resources: A.C.S., R.G.C., A.B., C.M.H.; Supervision: B.K.F., C.A.J., C.M.H.; Visualization: E.D.C.; Writing – original draft: E.D.C., C.M.H.; Writing – review & editing: E.D.C., M.Q., A.A., M.A.K., H.C., B.M., C.T., A.C.S., H.L.K., B.K.F., C.A.J., C.M.H.

## ETHICS DECLARATION

The MyCode Community Health Initiative is approved by the Institutional Review Board, and informed consent was obtained from all participants.

## REFERENCES

1. Corrado, D. & Zorzi, A. Filamin C: A New Arrhythmogenic Cardiomyopathy–Causing Gene? JACC Clin. Electrophysiol. 4, 515–517 (2018).

2. Ortiz-Genga, M. F., Cuenca, S., Dal Ferro, M., et al. Truncating FLNC Mutations Are Associated With High-Risk Dilated and Arrhythmogenic Cardiomyopathies. J. Am. Coll. Cardiol. 68, 2440–2451 (2016).

3. Begay, R. L., Graw, S. L., Sinagra, G., et al. Filamin C Truncation Mutations Are Associated With Arrhythmogenic Dilated Cardiomyopathy and Changes in the Cell–Cell Adhesion Structures. JACC Clin. Electrophysiol. 4, 504–514 (2018).

4. Brun, F., Gigli, M., Graw, S. L., et al. FLNC truncations cause arrhythmogenic right ventricular cardiomyopathy. J. Med. Genet. jmedgenet-2019-106394 (2020) doi:10.1136/jmedgenet-2019-106394.

5. Ader, F., De Groote, P., Réant, P., et al. FLNC pathogenic variants in patients with cardiomyopathies: Prevalence and genotype-phenotype correlations. Clin. Genet. 96, 317–329 (2019).

6. Hall, C. L., Akhtar, M. M., Sabater-Molina, M., et al. Filamin C variants are associated with a distinctive clinical and immunohistochemical arrhythmogenic cardiomyopathy phenotype. Int. J. Cardiol. 307, 101–108 (2020).

7. Oz, S., Yonath, H., Visochyk, L., et al. Reduction in Filamin C transcript is associated with arrhythmogenic cardiomyopathy in Ashkenazi Jews. Int. J. Cardiol. 317, 133–138 (2020).

8. Monda, E., Frisso, G., Rubino, M., et al. Potential role of imaging markers in predicting future disease expression of arrhythmogenic cardiomyopathy. Future Cardiol. fca-2020-0107 (2020) doi:10.2217/fca-2020-0107.

9. Sveinbjornsson, G., Olafsdottir, E. F., Thorolfsdottir, R. B., et al. Variants in NKX2-5 and FLNC Cause Dilated Cardiomyopathy and Sudden Cardiac Death. Circ. Genomic Precis. Med. 11, e002151 (2018).

10. Akhtar, M. M., Lorenzini, M., Pavlou, M., et al. Association of Left Ventricular Systolic Dysfunction among Carriers of Truncating Variants in Filamin C with Frequent Ventricular Arrhythmia and End-stage Heart Failure. JAMA Cardiol. 1–11 (2021) doi:10.1001/jamacardio.2021.1106.

11. Miller, D. T., Lee, K., Chung, W. K., et al. ACMG SF v3.0 list for reporting of secondary findings in clinical exome and genome sequencing: a policy statement of the American College of Medical Genetics and Genomics (ACMG). Genet. Med. (2021) doi:10.1038/s41436-021-01172-3.

12. Carey, D. J., Fetterolf, S. N., Davis, F. D., et al. The Geisinger MyCode community health initiative: an electronic health record-linked biobank for precision medicine research. Genet. Med. 18, 906–13 (2016).

13. Dewey, F. E., Gusarova, V., O’Dushlaine, C., et al. Inactivating variants in ANGPTL4 and risk of coronary artery disease. N. Engl. J. Med. 374, 1123–1133 (2016).

14. Staples, J., Maxwell, E. K., Gosalia, N., et al. Profiling and Leveraging Relatedness in a Precision Medicine Cohort of 92,455 Exomes. Am. J. Hum. Genet. 102, 874–889 (2018).

15. Carruth, E. D., Young, W., Beer, D., et al. Prevalence and Electronic Health Record-Based Phenotype of Loss-of-Function Genetic Variants in Arrhythmogenic Right Ventricular Cardiomyopathy-Associated Genes. Circ. Genomic Precis. Med. 12, 487–494 (2019).

16. Lin, M. F., Rodeh, O., Penn, J., et al. GLnexus: joint variant calling for large cohort sequencing. bioRxiv 343970 (2018) doi:10.1101/343970.

17. McLaren, W., Gil, L., Hunt, S. E., et al. The Ensembl Variant Effect Predictor. Genome Biol. 17, 122 (2016).

18. Jing, L., Ulloa Cerna, A. E., Good, C. W., et al. A Machine Learning Approach to Management of Heart Failure Populations. JACC Hear. Fail. (2020) doi:10.1016/j.jchf.2020.01.012.

19. Raghunath, S., Pfeifer, J. M., Cerna, A. E. U., et al. Deep Neural Networks can Predict New-Onset Atrial Fibrillation from the 12-lead Electrocardiogram and Help Identify Those at Risk of AF-Related Stroke. Circulation (In Press), (2021).

20. Carruth, E. D., Beer, D., Alsaid, A., et al. Clinical Findings and Diagnostic Yield of Arrhythmogenic Cardiomyopathy through Genomic Screening of Pathogenic or Likely Pathogenic Desmosome Gene Variants. Circ. Genomic Precis. Med. 14, 201–212 (2021).

21. Marcus, F. I., McKenna, W. J., Sherrill, D., et al. Diagnosis of arrhythmogenic right ventricular cardiomyopathy/dysplasia. Circulation 121, 1533–1541 (2010).

22. Towbin, J. A., McKenna, W. J., Abrams, D. J., et al. 2019 HRS expert consensus statement on evaluation, risk stratification, and management of arrhythmogenic cardiomyopathy. Hear. Rhythm 16, e301–e372 (2019).

23. Firth, D. Bias Reduction of Maximum Likelihood Estimates. Biometrika 80, 27 (1993).

24. Benjamini, Y. & Hochberg, Y. Controlling the false discovery rate: a practical and powerful approach to multiple testing. Journal of the Royal Statistical Society. Series B (Methodological) vol. 57 289–300 (1995).

25. Povysil, G., Chazara, O., Carss, K. J., et al. Assessing the Role of Rare Genetic Variation in Patients With Heart Failure. JAMA Cardiol. (2020) doi:10.1001/jamacardio.2020.6500.

26. Augusto, J. B., Eiros, R., Nakou, E., et al. Dilated cardiomyopathy and arrhythmogenic left ventricular cardiomyopathy: a comprehensive genotype-imaging phenotype study. Eur. Hear. J. - Cardiovasc. Imaging 44, 1–11 (2019).

27. Gigli, M., Stolfo, D., Graw, S., et al. Phenotypic Expression, Natural History and Risk Stratification of Cardiomyopathy Caused by Filamin C Truncating Variants. Circulation (2021) doi:10.1161/CIRCULATIONAHA.121.053521.

28. Gigli, M., Merlo, M., Graw, S. L., et al. Genetic Risk of Arrhythmic Phenotypes in Patients With Dilated Cardiomyopathy. J. Am. Coll. Cardiol. 74, 1480–1490 (2019).

29. Haggerty, C. M., Damrauer, S. M., Levin, M. G., et al. Genomics-First Evaluation of Heart Disease Associated With Titin-Truncating Variants. Circulation 140, 42–54 (2019).

30. Begg, C. B. On the use of familial aggregation in population-based case probands for calculating penetrance. J. Natl. Cancer Inst. 94, 1221–1226 (2002).

31. Choi, S. H., Jurgens, S. J., Weng, L. C., et al. Monogenic and polygenic contributions to atrial fibrillation risk results from a national biobank. Circ. Res. 200–209 (2020) doi:10.1161/CIRCRESAHA.119.315686.

32. Ware, J. S. & Cook, S. A. Role of titin in cardiomyopathy: from DNA variants to patient stratification. Nat. Rev. Cardiol. (2017) doi:10.1038/nrcardio.2017.190.

33. Sen-Chowdhry, S., Syrris, P., Prasad, S. K., et al. Left-Dominant Arrhythmogenic Cardiomyopathy. An Under-Recognized Clinical Entity. J. Am. Coll. Cardiol. 52, 2175–2187 (2008).

34. Corrado, D., Marra, M. P., Zorzi, A., et al. Diagnosis of arrhythmogenic cardiomyopathy: The Padua criteria. Int. J. Cardiol. 108653 (2020) doi:10.1016/j.ijcard.2020.06.005.

35. Casella, M., Gasperetti, A., Sicuso, R., et al. Characteristics of Patients with Arrhythmogenic Left Ventricular Cardiomyopathy: Combining Genetic and Histopathologic Findings. Circ. Arrhythmia Electrophysiol. (2020) doi:10.1161/CIRCEP.120.009005.

36. Smith, E. D., Lakdawala, N. K., Papoutsidakis, N., et al. Desmoplakin Cardiomyopathy, a Fibrotic and Inflammatory Form of Cardiomyopathy Distinct From Typical Dilated or Arrhythmogenic Right Ventricular Cardiomyopathy. Circulation 141, 1872–1884 (2020).

