## Supplemental Material for "Loss-of-Function *FLNC* Variants are Associated with Arrhythmogenic Cardiomyopathy Phenotypes when Identified through Exome Sequencing of a General Clinical Population"

**Supplementary Table 1.** Phenotype definitions

| Phenotype | Diagnosis Codes (ICD-10†) or Definition |
|---|---|
| Diagnosis codes in two or more encounters, or on the individual's Problem List: | |
| DCM | I42.0, I42.8, I42.9 |
| VT | I47.0, I47.2 |
| SVT | I47.1, I49.2 |
| PVCs | I49.3, I49.4 |
| ICD | Z95.810 |
| Diagnosis codes in one or more encounters, or on the individual's Problem List: | |
| Cardiac arrest | I46.2, I46.9 |
| VF | I49.01, I49.02 |
| Other phenotype definitions: | |
| AF | See Supplement from Raghunath et al. (2021)[1] |
| HF | Any of the following 3 cases:<br>1. Active HF diagnosis on problem list<br>2. eMERGE ("definite") phenotype<br>3. (≥2 outpatient encounter diagnoses OR ≥1 inpatient/emergency encounter diagnosis) AND HF medication prescription<br>HF diagnosis codes included:<br>I50.1, I50.2*, I50.3*, I50.4*, I50.9, I11.0, I13,0, I13.2, 428.0, 428.1, 428.2*, 428.3*<br>HF medications included:<br>Furosemide, Lasix, Bumetanide, Bumex, Torsemide, Demadex, Ethacrynic acid, Edecrin, Metolazone, Zaroxolyn |

ICD-10: international classification of diseases, tenth revision DCM: dilated cardiomyopathy; VT: ventricular tachycardia; SVT: supraventricular tachycardia; PVCs: premature ventricular contractions; ICD: implantable cardioverter-defibrillator; VF: ventricular fibrillation; AFib: atrial fibrillation; HF: heart failure. †Also includes mapped codes to other related code systems, including ICD-9.

**Supplementary Table 2.** List of cardiomyopathy- or arrhythmia-associated genes excluded from 1-to-1 matched control group

| **Gene List** | | | |
|---|---|---|---|
| *ABCC9* | *DSP* | *LDB3* | *RIT1* |
| *ACTC1* | *DTNA* | *LMNA* | *RYR2* |
| *ACTN2* | *EYA4* | *MAP2K1* | *SCN10A* |
| *AGL* | *FHL2* | *MAP2K2* | *SCN1B* |
| *AKAP9* | *FKRP* | *MIB1* | *SCN2B* |
| *ALMS1* | *FKTN* | *MYBPC3* | *SCN3B* |
| *ALPK3* | *FLNC* | *MYH6* | *SCN4B* |
| *ANK2* | *FXN* | *MYH7* | *SCN5A* |
| *ANKRD1* | *GAA* | *MYL2* | *SGCD* |
| *BAG3* | *GATAD1* | *MYL3* | *SLC22A5* |
| *BRAF* | *GPD1L* | *MYL4* | *SNTA1* |
| *CACNA1C* | *HCN4* | *MYLK2* | *SOS1* |
| *CACNA2D1* | *HRAS* | *MYOM1* | *TBX20* |
| *CACNB2* | *ILK* | *MYOZ2* | *TBX5* |
| *CALM1* | *JPH2* | *MYPN* | *TCAP* |
| *CALM2* | *JUP* | *NEBL* | *TGFB3* |
| *CALM3* | *KCNA5* | *NEXN* | *TMEM43* |
| *CASQ2* | *KCND3* | *NKX2-5* | *TMPO* |
| *CAV3* | *KCNE1* | *NRAS* | *TNNC1* |
| *CAVIN4* | *KCNE2* | *PDLIM3* | *TNNI3* |
| *CHRM2* | *KCNE3* | *PKP2* | *TNNT2* |
| *CRYAB* | *KCNH2* | *PLN* | *TPM1* |
| *CSRP3* | *KCNJ2* | *PRDM16* | *TRDN* |
| *DES* | *KCNJ5* | *PRKAG2* | *TRPM4* |
| *DMD* | *KCNJ8* | *PTPN11* | *TTN* |
| *DOLK* | *KCNQ1* | *RAF1* | *TTR* |
| *DSC2* | *KRAS* | *RANGRF* | *TXNRD2* |
| *DSG2* | *LAMA4* | *RBM20* | *VCL* |

Only individuals with rare missense or putative loss-of-function variants in the listed genes were excluded.

### Supplemental References

1. Raghunath, S., Pfeifer, J. M., Cerna, A. E. U., *et al.* Deep Neural Networks can Predict New-Onset Atrial Fibrillation from the 12-lead Electrocardiogram and Help Identify Those at Risk of AF-Related Stroke. *Circulation* **(In Press)**, (2021).
